# Continued Effectiveness of COVID-19 Vaccination among Urban Healthcare Workers during Delta Variant Predominance

**DOI:** 10.1101/2021.11.15.21265753

**Authors:** Fan-Yun Lan, Amalia Sidossis, Eirini Iliaki, Jane Buley, Neetha Nathan, Lou Ann Bruno-Murtha, Stefanos N. Kales

**Author notes:** Corresponding author: Stefanos N. Kales, MD, MPH, Occupational Medicine, Cambridge Health Alliance, Macht Building 427, 1493 Cambridge Street, Cambridge, MA 02139, Tel. 617/665-1580 Fax. 617/665-1672.

## Abstract

**Background:** Data on COVID-19 vaccine effectiveness (VE) among healthcare workers (HCWs) during periods of delta variant predominance are limited.

**Methods:** We followed a population of urban Massachusetts HCWs (45% non-White) subject to epidemiologic surveillance. We accounted for covariates such as demographics and community background infection incidence, as well as information bias regarding COVID-19 diagnosis and vaccination status.

**Results and Discussion:** During the study period (December 16, 2020 to September 30, 2021), 4615 HCWs contributed to a total of 1,152,486 person-days at risk (excluding 309 HCWs with prior infection) and had a COVID-19 incidence rate of 5.2/10,000 (114 infections out of 219,842 person-days) for unvaccinated person-days and 0.6/10,000 (49 infections out of 830,084 person-days) for fully vaccinated person-days, resulting in an adjusted VE of 82.3% (95% CI: 75.1–87.4%). For the secondary analysis limited to the period of delta variant predominance in Massachusetts (i.e., July 1 to September 30, 2021), we observed an adjusted VE of 76.5% (95% CI: 40.9–90.6%). Independently, we found no re-infection among those with prior COVID-19, contributing to 74,557 re-infection-free person-days, adding to the evidence base for the robustness of naturally acquired immunity.

## Background

Data on COVID-19 vaccine effectiveness (VE) among healthcare workers (HCWs) during periods of delta variant predominance are limited. Literature accounting for other potential determinants of infection rates is even more scarce.

## Objective

To investigate the continued effectiveness of COVID-19 vaccination during the Delta variant predominance in a diverse and urban healthcare setting.

## Methods and Findings

A community-based healthcare system in Massachusetts runs a COVID-19 vaccination program for employees (described previously (1)), with the Pfizer vaccine starting on December 16, Moderna on December 23, 2020, and J&J/Janssen in February 2021. Vaccination was available to all workers regardless of their in-person/remote working status from December 29, 2020. In addition, the system announced a vaccine mandate on August 16, 2021, which requires employees to receive their final dose by October 18, 2021 barring an approved religious or medical exemption.

We followed all actively serving HCWs in the system from December 16, 2020 to September 30, 2021, excluding those with prior COVID-19 infection from the main analyses. The outcome was having a positive PCR assay during the study period documented by the healthcare system’s Occupational Health department (2). The established master database, comprised of workers’ demographics, prior infection, *de novo* PCR positivity, vaccination (validated by the Massachusetts Immunization Information System and/or the healthcare system’s medical records), and human resource administrative data, has been previously described (1, 2). For each HCW, we calculated the person-days at risk and categorized them according to vaccination status. A HCW’s follow-up person-days were censored at the end of the study period, his/her termination date, the date tested positive for COVID, or the date he/she received a 3rd vaccine dose, whichever came first.

The Andersen-Gill extension of the Cox proportional hazards models were built to account for correlated data. We further adjusted for age, sex, race, and the Massachusetts statewide 7-day average of tested COVID cases (3) on the date the first dose was given to control for background rates. We estimated VE by calculating 100%×(1−hazard ratio). The R software (version 3.6.3) was used for statistical analyses.

A total of 4,615 HCWs (average age of 45.0±13.3 years and female predominance (76.0%)) contributed to 1,152,486 person-days at risk during the study period. Forty-five percent of the study population was non-White (including 20% African American, 13.5% Hispanic, and 9.0% Asian). Of all HCWs, 4,418 (95.7%) had received at least one dose by the end of the study. Among them, 58.3% got Moderna, 39.4% Pfizer, 2.3% J&J/Janssen, and one (0.02%) got mixed doses of J&J/Janssen and Moderna. The results showed that throughout the study period, for fully vaccinated HCWs the VE is 82.3% (95% CI: 75.1–87.4%) after multivariable adjustment (Table, Figure).

**Table.**
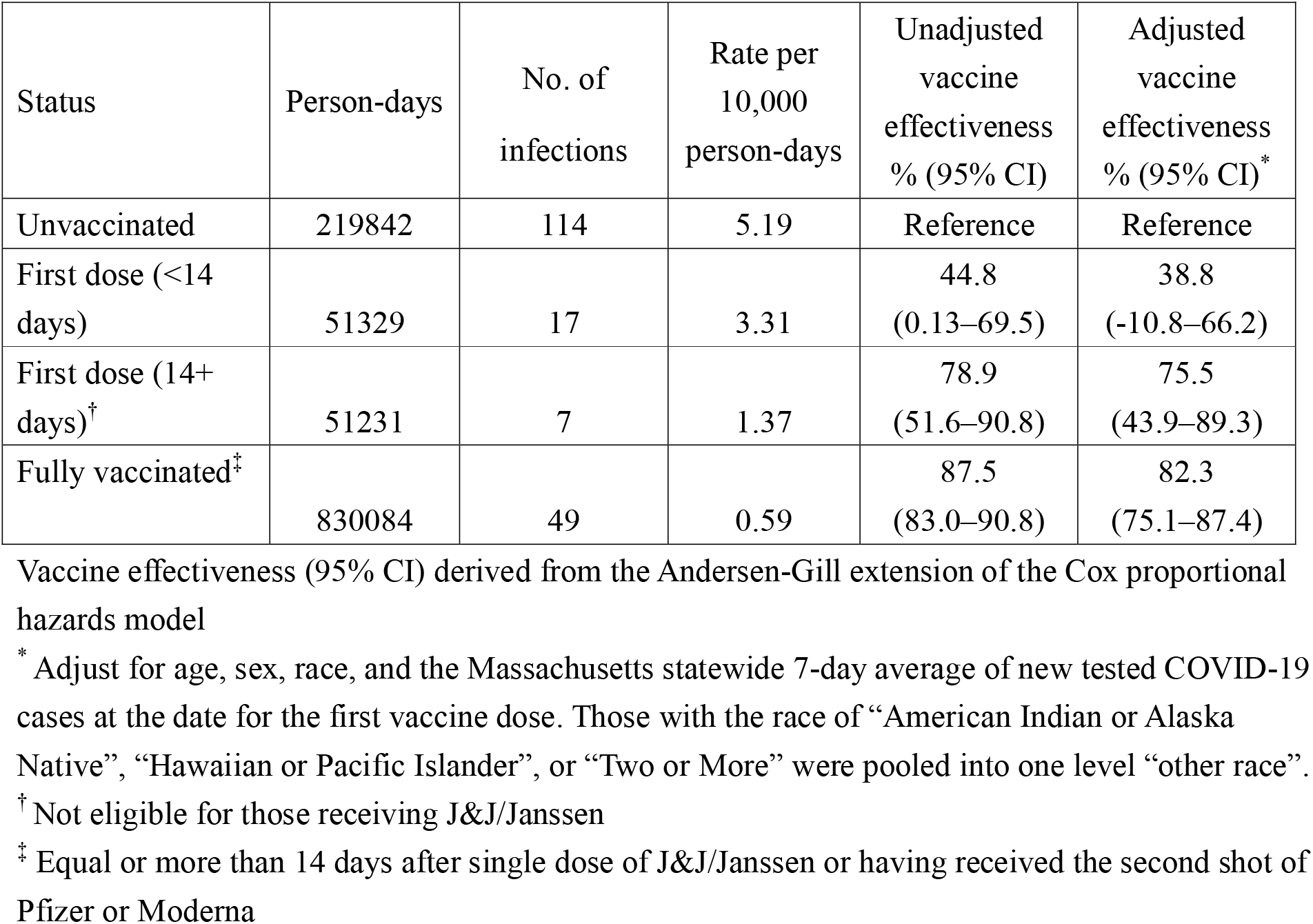
Rate of infection during the study period (Dec 16, 2020 – Sep 30, 2021) across the four vaccination categories (excluding 309 people infected before Dec 15, 2020)

| Status | Person-days | No. of infections | Rate per 10,000 person-days | Unadjusted vaccine effectiveness % (95% CI) | Adjusted vaccine effectiveness % (95% CI)* |
| --- | --- | --- | --- | --- | --- |
| Unvaccinated | 219842 | 114 | 5.19 | Reference | Reference |
| First dose (<14 days) | 51329 | 17 | 3.31 | 44.8<br>(0.13–69.5) | 38.8<br>(-10.8–66.2) |
| First dose (14+ days)<sup>†</sup> | 51231 | 7 | 1.37 | 78.9<br>(51.6–90.8) | 75.5<br>(43.9–89.3) |
| Fully vaccinated<sup>‡</sup> | 830084 | 49 | 0.59 | 87.5<br>(83.0–90.8) | 82.3<br>(75.1–87.4) |

Vaccine effectiveness (95% CI) derived from the Andersen-Gill extension of the Cox proportional hazards model
\* Adjust for age, sex, race, and the Massachusetts statewide 7-day average of new tested COVID-19 cases at the date for the first vaccine dose. Those with the race of “American Indian or Alaska Native”, “Hawaiian or Pacific Islander”, or “Two or More” were pooled into one level “other race”.
<sup>†</sup> Not eligible for those receiving J&J/Janssen
<sup>‡</sup> Equal or more than 14 days after single dose of J&J/Janssen or having received the second shot of Pfizer or Moderna

**Figure.**
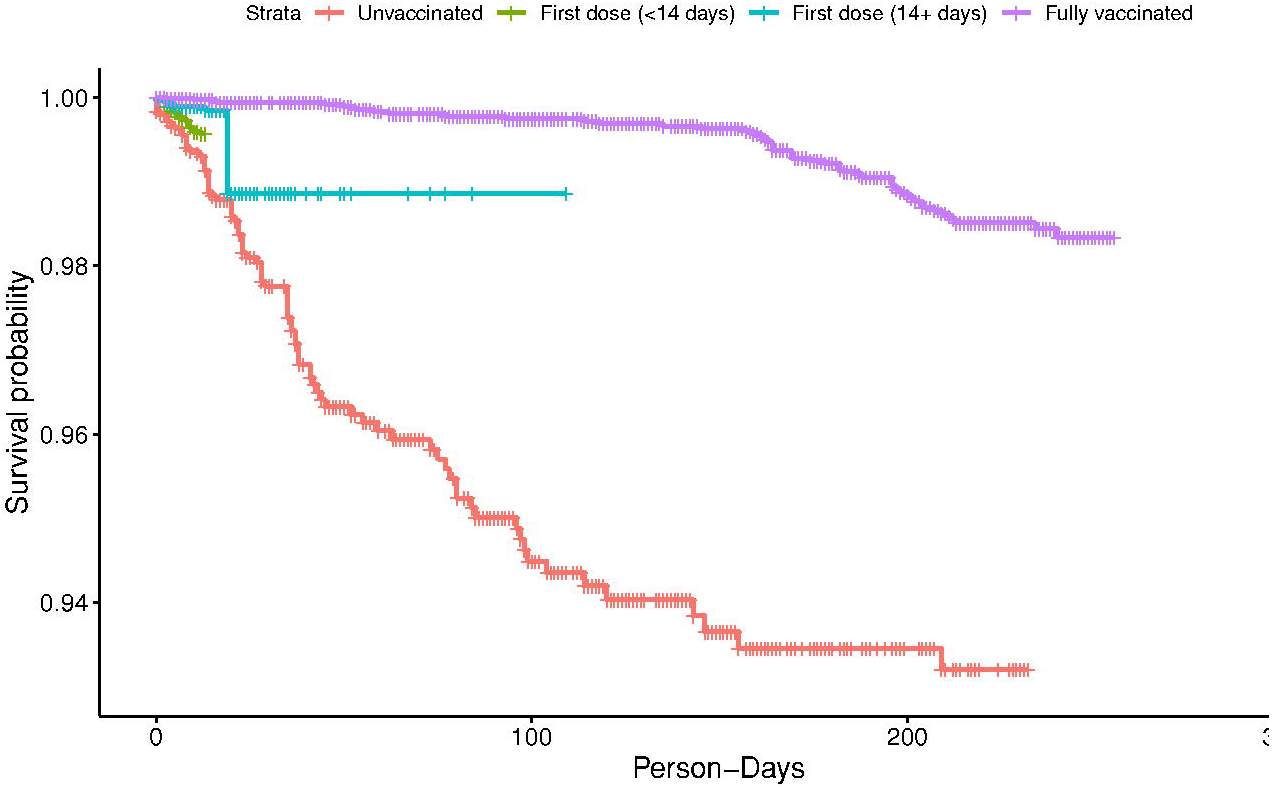
The Kaplan-Meier curve for the survival (i.e. infection-free) person-days across the four categories based on vaccination status.

We further conducted a secondary analysis limiting the study period from July 1, 2021 to September 30, 2021, corresponding to delta variant predominance in Massachusetts (4). We observed an incidence rate of 5.8/10,000 (15 events out of 25,910 person-days) for unvaccinated person-days and 1.3/10,000 (39 events out of 308,267 person-days) for 14 days after fully vaccinated, resulting in an adjusted VE of 76.5% (95% CI: 40.9–90.6%).

When we examined HCWs (n=423) with infections occurring before vaccination, no re-infection was observed, accumulating 74,557 re-infection-free person-days (starting 10 days after initial infection and censoring at the date of receiving their first vaccine dose). Further, after vaccination, previously infected HCWs did not contribute any breakthrough infection events among the vaccinated HCWs.

## Discussion

To our knowledge, this study is one of the first in healthcare settings regarding continued VE during delta variant predominance. Our work also provides further evidence of naturally acquired immunity. We found similar VE against the delta variant, 76%, compared to another study’s findings, 66% (5). Strengths included accounting for covariates and information bias such as demographics and background incidence, a multiethnic study population, consistent COVID-19 screening criteria, and well-validated vaccination records. Nonetheless, we did not examine individual manufacturers’ VE due to a limited power.

## Data Availability

All data produced in the present study are available upon reasonable request to the authors.

## Notes

Conflict of interest disclosure: S.N.K. has received COVID-19-related consulting fees from Open Health and has owned shares of Regeneron, Moderna and Astra-Zeneca. All other authors declare no competing interests.

Financial support: None reported.

### Competing Interest Statement

S.N.K. has received COVID-19-related consulting fees from Open Health and has owned shares of Regeneron, Moderna and Astra-Zeneca. All other authors declare no competing interests.

### Funding Statement

This study did not receive any funding.

### Author Declarations

The data we used were de-identified, the need for a consent was waived and the study was exempted by the Cambridge Health Alliance Institutional Review Board. (Protocol number 4/29/202-003)

